# Cell-free DNA from ascites identifies clinically relevant variants and tumour evolution in patients with advanced ovarian cancer

**DOI:** 10.1101/2024.03.09.24303840

**Authors:** Bonnita Werner, Elyse Powell, Jennifer Duggan, Marilisa Cortesi, Yeh Chen Lee, Vivek Arora, Ramanand Athavale, Michael Dean, Kristina Warton, Caroline E. Ford

**Affiliations:** Gynaecological Cancer Research Group, School of Clinical Medicine, Faculty of Medicine and Health, University of New South Wales, Australia; Gynaecological Oncology Department, Royal Hospital for Women, Sydney, Australia; Laboratory of Cellular and Molecular Engineering, Department of Electrical, Electronic and Information Engineering, Alma Mater Studiorum- University of Bologna.; School of Clinical Medicine, Faculty of Medicine and Health, University of New South Wales, Sydney, Australia; Prince of Wales Private Hospital, Sydney, Australia; Laboratory of Translational Genomics, Division of Cancer Epidemiology and Genetics, National Cancer Institute, Maryland, USA

**Keywords:** cell-free DNA, ascites, ovarian cancer, precision medicine, biomarkers, homologous recombination deficiency, targeted medicine

## Abstract

**Background:** The emergence of targeted therapies and predictive biomarkers is transforming the ovarian cancer treatment paradigm. However, the demand for high quality, tumour-enriched samples for biomarker profiling can be limited by access to adequate tissue samples. The use of cell-free DNA (cfDNA) in ascites presents a potential solution to this clinical challenge.

**Methods:** A unique set of sequential ascites-derived cfDNA samples (26 samples from 15 human participants) were collected from people with ovarian cancer (age range 36-82 years). cfDNA was sequenced using targeted next-generation sequencing, along with matched DNA from ascites-derived tumour cells (n=5) and archived FFPE-tissue from surgery (n=5).

**Results:** Similar tumour purity, variant detection and reference alignment were achieved with cfDNA when compared to FFPE and ascites derived tumour cell DNA, as well as improved coverage. No artefactual single-base mutation signatures were identified in cfDNA. Combined analysis of large-scale genomic alterations, loss of heterozygosity and tumour mutation burden identified 6 cases of high genomic instability (including 4 with pathogenic variants in *BRCA1* and *BRCA2*). Copy number profiles and subclone prevalence changed between sequential ascites samples, particularly in a case study where deletions and chromothripsis in Chr17p13.1 and Chr8q resulted in changes in clinically relevant *TP53* and *MYC* variants over time.

**Conclusions:** Ascites cfDNA successfully identified clinically actionable information, concordant to tissue biopsies, enabling opportunistic molecular profiling. These findings advocate for analysis of ascites cfDNA *in lieu* of accessing tumour tissue via biopsy.

## 1. Introduction

The rise of targeted agents has seen a pivotal shift in ovarian cancer treatment ^1^. Consequently, individualised biomarker profiling is now standard clinical practice^2^. Biomarker identification is reliant on accessible, tumour enriched DNA sources, which can be a challenge, particularly in circumstances where excisional or core biopsies are contraindicated or intolerable to the patient ^3, 4^. Ascites is a common feature of advanced ovarian cancer and may provide access to informative, heterogenous tumour tissue *in lieu* of a tissue biopsy, avoiding the need for additional invasive procedures (Figure 1A).

**Figure 1.**
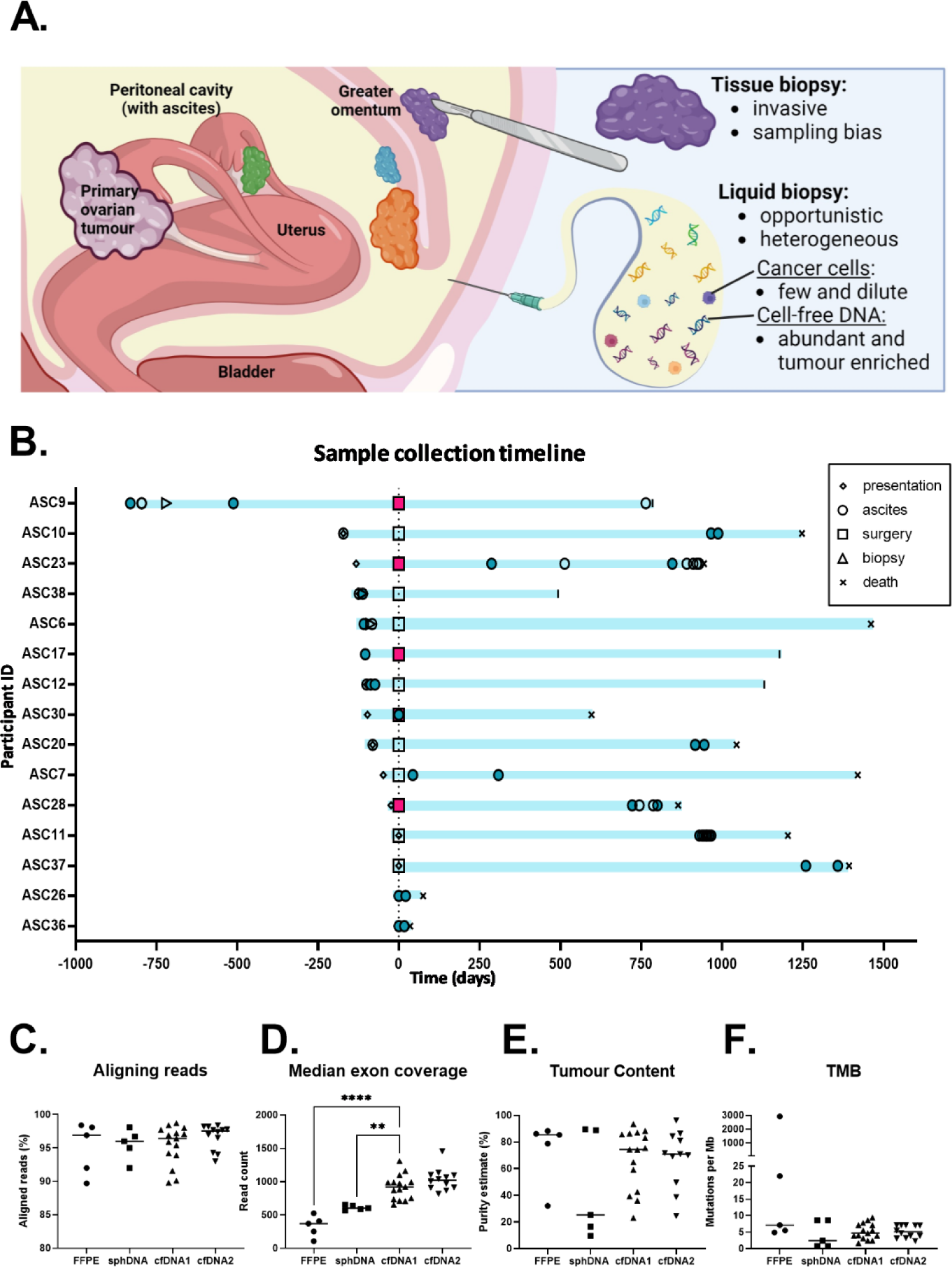
Sample collection overview and quality metrics. (A) Advantages and limitations of tissue and liquid biopsies. (B) Timeline of sample collection events with samples included in study as solid shapes. (C) Percentage of reads successfully aligning to GRCh37, (D) median read depth over targeted exons, (E) Tumour mutation burden (TMB) determined by TSO500 local app v2.2 and (F) Tumour purity estimated from deleterious TP53 VAF, compared by Dunnet’s multiple comparisons test (though serial cfDNA samples are visualised, only the first sample is included in analysis). cfDNA, cell-free DNA; sphDNA, DNA from ascites-derived cell spheroids; FFPE, DNA from

Ascites is present at the time of diagnosis in over 90% of stage III and IV ovarian cancers, where most diagnoses are made ^5, 6^, and is drained to relieve the debilitating symptoms it causes ^7^. The incidentally collected ascites samples are often used as surrogate biopsies for ovarian cancer, to aid diagnosis ^8^. However, approximately 20% of these specimens are cytologically classified as non-malignant due to absence of cancer cells, hindering performance of molecular tumour analysis ^9, 10^. Tumour enrichment of ascites cells can be accomplished by collecting only cell spheroids (the metastatic drivers of ovarian cancer) ^11^, however, we have previously shown cell-free DNA (cfDNA) to improve on spheroids as a source of concentrated and abundant tumour DNA ^12^. This is in alignment with other reports demonstrating that ascites-derived cfDNA is tumour enriched ^13^, even in cases where malignant cells are absent from the fluid ^10, 14^. Representative (even unique) mutational profiles have been proven identifiable in cfDNA and homologous recombination deficiency (HRD) scores that align with solid tumours have also been elucidated ^15-17^. Consequently, recommendations have been made to implement cfDNA testing in routine clinical practice. However, limited focus has been placed on verifying unique mutations in cfDNA.

As ascites poses a poor prognosis for primary debulking surgery ^18, 19^, neoadjuvant chemotherapy is often used ^20^, delaying access to tumour tissue for molecular profiling, or necessitating invasive and often unsuccessful core biopsies ^21, 22^. Tissue collected at interval debulking surgery often performs poorly in sequencing, due to chemotherapy causing the death of tumour cells and increasing immune infiltrate, diluting tumour DNA ^23^. Additionally, ascites occurs in most cases of relapse, where further surgery is rarely performed ^3^. Relapsed ovarian cancer is often molecularly distinct from its predecessor ^24, 25^. Thereby cfDNA in ascites may provide earlier access to an uncompromised tumour sample, as well as unique access to evolved disease. However, of studies on ovarian cancer ascites cfDNA, none have centred on sequential ascites samples for the temporal analysis of ovarian cancer.

In this study we aimed to understand whether new and useful information could be gained from ascites cfDNA at different time points, with a focus on whether this information is reliable and reproducible. We assess the feasibility of applying targeted next-generation sequencing to ascites cfDNA to identify actionable biomarkers, and we evaluate concordance with ascites tumour cells, archived formalin-fixed tumour tissue and clinical reports. We also compare cfDNA from sequential ascites samples to ascertain whether opportune ascites sampling reveals time-critical changes that may inform personalised disease management.

## 2. Methods

### 2.1 Cohort selection and sample collection

Ascites samples were collected with informed consent during routine paracentesis appointments, from patients of the Royal Hospital for Women, Sydney, with confirmed or suspected ovarian cancer. The 15 person cohort for this study (as part of a larger study ^12^) was selected based on meeting any of the ordered priorities when this study began: multiple ascites samples collected; clinical sequencing report available identifying deleterious *BRCA1/2* variants (germline or somatic); time-matched tissue sample available; clinical sequencing report available identifying deleterious somatic variants (Supplementary Figure 1). Where available, FFPE tumour biopsy samples were retrieved through the Health Systems Alliance Biobank, UNSW. Research IDs used are not known to anyone outside the research group. This study was conducted under approval from the Prince of Wales Hospital Human Research Ethics Committee (HC19- 001).

### 2.2 Sample processing and DNA extraction

Samples were processed to fractionate cell-free fluid and cell spheroid pellets, as previously described ^12^. Briefly, fluid was passed through a 40 µm filter, capturing cell spheroids. Filtrate underwent two centrifugations to isolate the ‘cell-free’ component. cfDNA was extracted from cell-free ascites fluid using the QIAamp Circulating Nucleic Acid Kit (Qiagen). DNA was extracted from cell spheroid pellets using the All-In-One DNA/RNA/Protein Miniprep kit (Astral Scientific). FFPE tissue was first treated with de- paraffanization solution (Qiagen), then DNA was extracted using the NucleoSpin DNA FFPE XS, Microkit for DNA from FFPE (Macherey-Nagel) or the QIAamp DNA FFPE Tissue Kit (Qiagen).

### 2.3 Next Generation Sequencing

TruSight Oncology 500 (TSO500) libraries, covering 523 cancer-related genes, were prepared at the Ramaciotti Centre for Genomics, UNSW. DNA integrity was assessed using the Agilent gDNA ScreenTape or the Cell-free DNA ScreenTape on the Agilent TapeStation 4200, for spheroid DNA (sphDNA) and cfDNA, respectively. FFPE DNA integrity was tested with the Illumina FFPE QC Kit.

Samples were prepared according to the Illumina TSO500 High Throughput Reference Guide, following the DNA only workflow, with a minimum of 40 ng input, measured by Qubit High Sensitivity DNA Assay (Thermo Fisher Scientific). sphDNA and FFPE samples were fragmented using the Covaris E220, then checked on the High Sensitivity D1000 ScreenTape assay. cfDNA samples were not fragmented.

Final libraries were checked on the High Sensitivity D1000 ScreenTape assay and bead-based normalisation was performed to ensure quality and uniformity prior to pooling. Library pools were prepared following the relevant instructions in the NovaSeq 6000 Denature and Dilute Libraries Guide. Samples were sequenced on the NovaSeq 6000 using either the S1 200-cycle kit or the SP 300-cycle kit (XP protocol), following manufacturer instruction.

Oxford nanopore technology (ONT) sequencing library for participant ASC23 was created using the ligation sequencing gDNA (SQK-LSK110) kit from an input of ∼30 ng cfDNA (determined by Qubit). Library preparation proceeded without multiplexing and with some modifications to the protocol to account for cfDNA’s fragment length, as recommended by ONT (https://community.nanoporetech.com/extraction_methods/human-blood-cfdna).

AMPure XP Bead (Beckman Coulter, Inc.) ratio was increased and MinKNOW settings were adjusted to allow capture of fragments as short as 20bps. An R9.4.1 flow cell sequenced 50 fmol of cfDNA library on a GridION with MinKNOW software. Epi2Me software was used to convert FAST5 to FASTQ files and align to GRCh38. ONT data was managed on the Seven Bridges Cancer Research Data Commons Cloud Resource ^26^. Reads per megabase (Mb) aligning to Chr17p13.1 (17:6,500,001-10,800,000) and 17q21.31 (42,800,001-46,800,000) were counted.

### 2.3 Data processing and analysis

Full details of data processing and analysis are listed in Supplementary File 1. Briefly, sequencing data was processed using the TSO500 v2.2 Local Application (TSO500 Local App), aligning to reference genome hg19/GRCh37 ^27^. The app reported tumour mutation burden (TMB) and gene-specific amplifications.

As we had no germline reference samples, variants were considered likely somatic if they had a minor allele count of less than 100 in each of three population databases (<0.05% GnomAD Exome, <0.5% GnomAD Genome and <2% 1000 Genomes). Of these, SNPs and MNPs were assessed by the Cancer Genome Interpreter (CGI) ^28^, using SNPnexus ^29^, to assess their likelihood of cancer driving capacity. ClinVar, Varsome and COSMIC were used to similarly assess insertions and deletions. Likely somatic variants’ single base substitution signature was identified using SigProfiler tools, SigProfiler Matrix Generator and SigProfiler Extractor, using the computational cluster Katana ^30^.

Tumour purity was estimated by the following formula, where *t* is tumour fraction and *v* is the variant allele frequency (VAF) of the deleterious *TP53* variant.

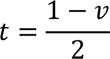

For patient ASC23, tumour purity was estimated based on proportionate deleterious *PIK3CA* variant VAF in cfDNA and sphDNA compared to FFPE, made relative to FFPE estimated tumour purity (based on *TP53*).

Copy number (CN) profiles were estimated using CNVKit ^31^ with Katana ^30^ and were clustered using the ‘Clustermap’ function of Seaborn Python package. Segments of disparate CN which were >10Mb in length, and adjacent to other >10Mb segments on the same chromosome, were considered large genome alterations (LGAs) ^32^. Loss of heterozygosity was identified by counting the percentage of occupied chromosome- limited 100KB bins with average VAF deviating from 0.4-0.6.

Cancer clone clusters were estimated using PyClone-VI ^33^. In one case (ASC28), the deleterious *TP53* variant could not be clustered, although considered a likely gatekeeper mutation (see Supplementary File 1), so a high frequency unrepresented clone was assumed, coloured grey. Cancer cell frequency of clones at different time points were used to estimate possible phylogeny and evolution.

### 2.4 Statistics

Non-parametric statistical tests, listed in associated figure legends, were performed using GraphPad Prism 9.5.1 software. Results were considered significant where p-value was below 0.05.

## 3. Results

### 3.1 Cohort

This study recruited 15 participants, 14 with high grade serous tubal/ovarian/peritoneal cancer and one with clear cell ovarian cancer. Median age at recruitment was 61 years (age range, 36-82 years). Seven of the 15 participants were recruited with chemotherapy naïve disease, one after minimal prior chemotherapy, one at interval debulking surgery and six with recurrent disease (with all but one of these having had previous chemotherapy, Figure 1B). Participant characteristics are listed in Supplementary Table 1.

### 3.2 cfDNA from ascites is an exemplary template for next-generation sequencing

#### 3.2.1 Alignment, coverage and tumour purity

Targeted sequencing was performed on cfDNA (n=26), FFPE (n=5) and sphDNA (n=5) samples. No significant difference in genome alignment was observed (ordinary one-way ANOVA, p=0.61, Figure 1C), but cfDNA achieved significantly higher median exon coverage (MEC, mean 915.9) compared to sphDNA (mean MEC 610.8, p=0.0057) and FFPE (mean MEC 332.6, p<0.0001) by ordinary one-way ANOVA and Dunnet’s multiple comparisons test (Figure 1D).

The tumour purity of cfDNA samples was estimated at 23%-93%, with 14 of 15 cfDNA samples having >30% tumour purity (Figure 1E). By comparison, estimated tumour purity ranged from 10%-90% in sphDNA, with 3 of 5 <30% pure, and from 32%-92% in FFPE. Comparing patient-matched samples, cfDNA had higher purity than sphDNA in 3 of 5 cases (range 0.25-5.03x purity, average 2.73±2.10SD) and higher purity than FFPE in 2 of 5 cases (range 0.29-1.21x, average 0.88±0.36, Supplementary Figure 2A).

The TMB of cfDNA (average 5.2±2.5SD) was not significantly different to FFPE (Dunnet’s multiple comparison’s test, p=0.10) or sphDNA (p>0.99, Figure 1D). Where comparing patient-matched samples, cfDNA identified 2.99±2.38SD times the TMB of sphDNA (range 1-6.88x) and 0.73±0.26SD times the TMB of FFPE (considering only the three samples where artefacts were not detected, range 0.49-1x, Supplementary Figure 2B).

#### 3.2.2 Identification of key clinical variants

Somatic cancer driving variants were detected in all cfDNA samples (3-8 deleterious/VUS variants per patient) at VAFs of up to 0.88. Across the cohort, commonly altered genes included *TP53* (14 of 15), *BRCA1* (4 of 15, including 3 germline), *BRCA2* (3 of 15, including 1 germline), *NF1* (3 of 15), *RB1* (3 of 15) and *NOTCH1* (3 of 15)(Figure 2A, Supplementary Figure 3). Of these, all but *NOTCH1* are among the top 9 significantly recurrent genes identified in the TCGA ovarian carcinoma cohort ^34^.

**Figure 2.**
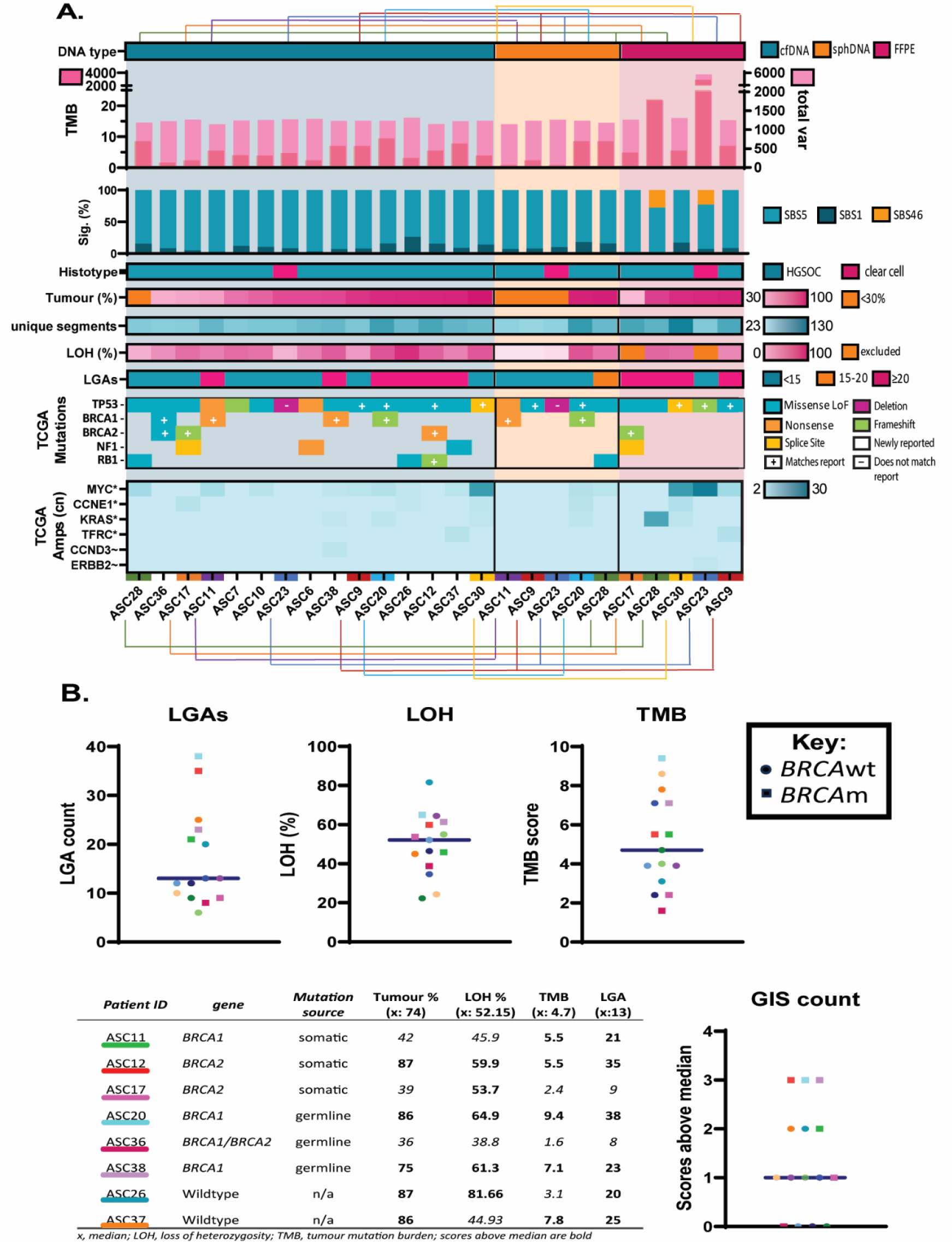
Sample characteristics, variant detection and genomic instability. (A) TSO500 output from initial cfDNA samples, sphDNA and FFPE, separated into DNA type, and arranged by increasing estimated tumour proportion. TMB – somatic variants per megabase, overlayed over total small variants detected (inclusive of germline and somatic) on the right Y-axis; Sig (%) – proportion of variants assigned to COSMIC single base substitution (SBS) signatures (sig); Histotype – high grade serous (HGSOC) or clear cell histological subtype; Tumour (%) – estimated tumour proportion >30%; unique segments – the number of genome segments with copy numbers unique to their surrounding sequences; LOH(%) – percentage of 100kb bins where non-homozygous variants (germline and somatic, 0.05<VAF<0.95) are, on average, outside of the expected range for heterozygosity (0.4-0.6 VAF), ie. with loss of heterozygosity (LOH), excluding samples with average read depth <150x or median read depth <0.1; Large-scale genomic alterations (LGAs) – number of >10MB segments with altered copy number and adjacent to other >10MB segments; Mutations – deleterious variants identified in genes known to be frequently mutated in ovarian cancer; amplifications – estimated copy number in genes known to be frequently amplified in ovarian cancer. (B) Number of large-scale genomic alterations (LGAs), percent of occupied 100KB genome bins with loss of heterozygosity (LOH) and tumour mutation burden (TMB) in initial cell-free DNA samples, with median indicated. Number of scores above the indicated median (GIS count). Colours are assigned to individuals and are maintained across A-D. cfDNA, cell-free DNA; sphDNA, DNA from ascites-derived cell spheroids; FFPE, DNA from formaldehyde-fixed paraffin embedded tumour biopsy samples; total var, total small variants reported; LoF, loss of function; wt, wildtype; m, mutant.

Where results of clinical testing were available (11 of 15 cases), concordance was seen in cfDNA for 18 of 19 clinically reported variants (Supplementary Table 2). The single mutation which was not identified in cfDNA was accounted for by a gene deletion event, discussed further in section 3.3.2.

Gene amplifications were identified in 87% of cfDNA samples (13 of 15, Figure 2A). The most commonly amplified genes included *MYC* (n=9) and *PIK3CA* (n=6), with other amplifications of potential clinical significance in *CCNE1* (n=3) and *EGFR* (n=2).

LOH, LGA and TMB were used as surrogate markers for genomic instability (Figure 2B). The three samples that scored above the median across all markers had *BRCA* mutations. Two additional *BRCA* wild-type samples (with no other pathogenic variants in known HRD-related genes) also scored highly, potentially harbouring *BRCA1* promoter methylation or another cause of HRD. Two *BRCA* mutant samples did not score highly across the markers, though these samples had lower tumour purity (36-39% vs. 75-87%). 100% of ascites samples had LOH in >16% of the assessed genome, the cut-off used by Coleman *et al.* (2017), though this is likely because targeted, as opposed to whole genome sequencing was used ^35^.

#### 3.2.3 Variant reporting accuracy and concordance across biospecimens

All cfDNA and sphDNA samples aligned to varying proportions of SBS1 and SBS5, both associated with ovarian cancer (Figure 2A, Supplementary Figure 4). The two FFPE samples with outlying number of variants had >20% alignment to artefactual signature SBS46, indicative of possible sequencing artefact. These two samples also had the highest reported TMB and an outlying number of total variants, hindering confident identification of real mutations.

Identified deleterious variants were concordant across most patient-matched samples, despite the fact that FFPE was collected at a different timepoint in most cases (Supplementary Figure 3). However, up to 7 unique variants were identified in cfDNA samples (including driver mutations in *KIT* and *NOTCH1,* 0.03 and 0.05VAF respectively, Supplementary Figure 5). Of the 2 participants in whom there was sufficient quality to verify variants in FFPE, one had 90% consensus (54 of 60) across all samples but the other had 10 (of 52) variants unique to FFPE. Among these, 8 were within a 3.2kb span on 6q22.1 (within gene *ROS1*), with an average VAF of 0.06. The unique variants were identified to belong to a single clone, isolated to the FFPE sample.

In other cases, sample sets had similar clonal representation. However, unique clones were reported in ascites samples in 3 cases (2 in cell-free DNA and 1 in sphDNA). Unique clones were also identified in FFPE in 5 participants (including 2 without confounding circumstances: no artefactual variants and adequate read depth over all samples).

CN variations were identified in all cfDNA samples (Figure 2A, Supplementary Figure 6). These CN profiles accurately clustered patient-matched sequential cfDNA and sphDNA samples together in all but one case, however only 2 of 5 FFPE samples correctly clustered with their patient-matched samples (Supplementary Figure 6).

In a case study comparing duplicate cfDNA sequencing reports from a single ascites sample, tumour fraction, CNV segmentation and LOH all varied by only 3% (93.48% vs 96.28%, 74 vs 76 and 64.46% vs 62.28%, respectively), LGA varied by 1 (13 vs 14) and TMB was identical (3.9).

### 3.3 Temporal change is evident with opportunistic ascites cfDNA sampling

#### 3.3.1 Newly diagnosed versus recurrent disease

To investigate a temporal effect in participants recruited at different stages of a typical ovarian cancer disease trajectory, we compared GIS markers in initial cfDNA samples from participants recruited before and after chemotherapy exposure.

We found no significant change in participants between chemotherapy status (Supplementary Figure 7), though a trend towards increased TMB was observed with chemotherapy exposure.

In the 6 participants with *BRCA1/BRCA2* mutations, we saw no evidence of reversion mutations in the mutated gene.

#### 3.3.2 Change between sequential ascites samples

Sequential ascites samples were collected from 11 participants, with intervals ranging from 13-559 days (average 132.5). We compared the paired cfDNA samples to consider both reproducibility and temporal changes.

We observed changes in CN profile over time (Figure 3A), with disparity in 6.78-48.75% of sites (average 20.07±11.44SD%). Though not significant, participants with more extensive chemotherapy history between samples tended to have larger divergence between samples (note participant ASC28 had tumour content below the recommended cut-off of 30% for the CNV profiling, likely contributing to overestimation of CNV). Among clones identified in each participant, we identified 5 cases where CCF in at least one cluster altered by >10% (Figure 3B). No sample pairs had clones unique to one sample.

**Figure 3.**
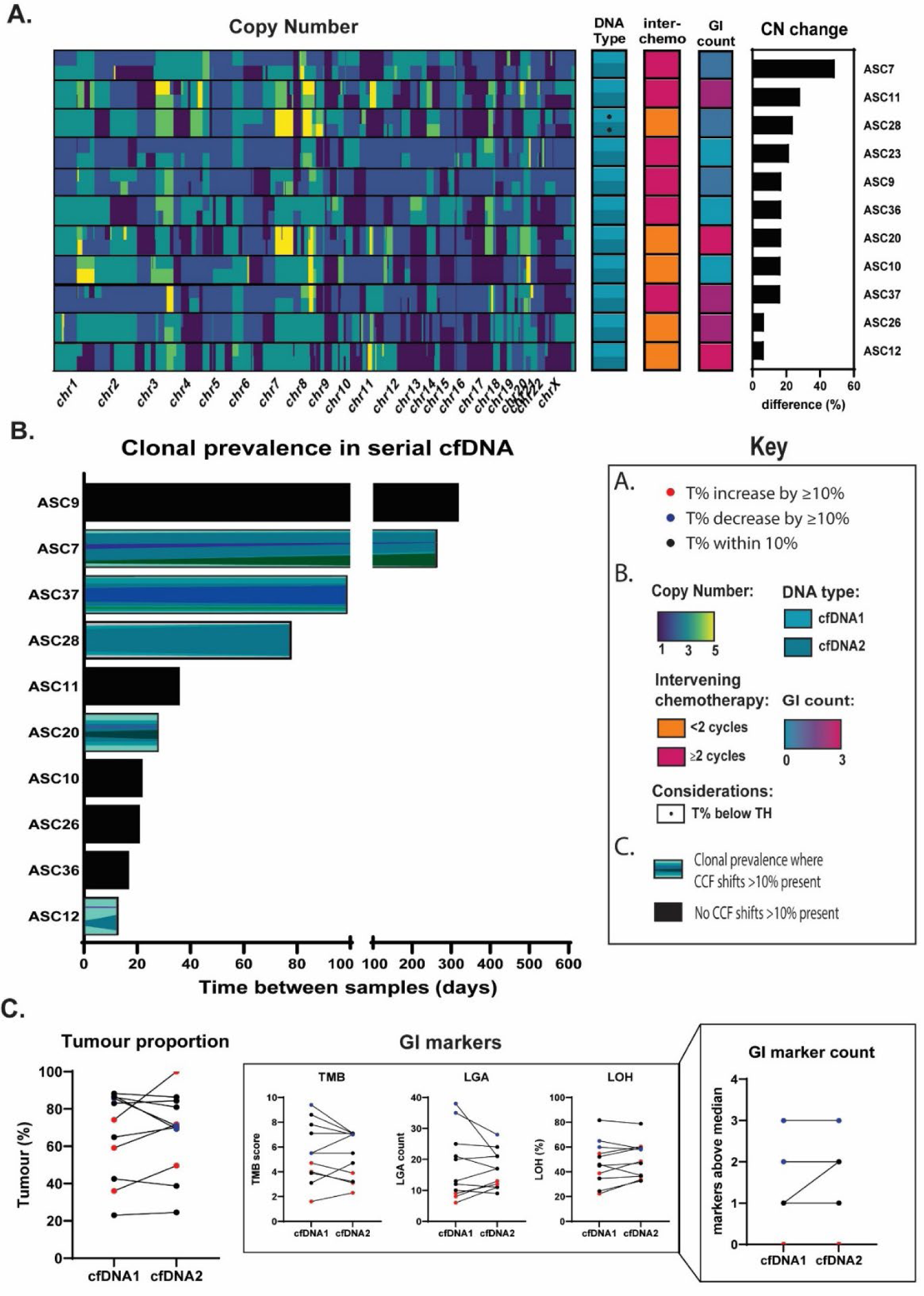
Temporal change in ascites cfDNA. (A) Copy number profile of serial cfDNA samples, and calculated percentage of queried sites with disparate copy number profiles, in association with intervening chemotherapy history. (B) Representations of clonal cancer cell fractions in serial ascites samples where there are fraction changes >10% between samples. (C) Markers of genomic instability (tumour mutation burden, large-scale genomic alterations and loss of heterozygosity) in serial ascites samples, with sample pairs which varied in tumour proportion by more than 10% marked in red (increase) or blue (decrease) according to left-most figure. Right-most figure indicates number of markers above threshold for each individual.cfDNA, cell-free DNA; T%, tumour percentage; TH, threshold of 30% tumour recommended for copy number analysis; chr, chromosome.

We did not observe any instances of driver mutation emergence or disappearance between cfDNA pairs. We also did not observe any instances of unique gene amplifications (greater than 2-fold). Though we did see some changes in genome instability markers (Figure 3C), we did not identify any significant trends associated with amount of time/chemotherapy between samples (Supplementary Figure 8). However, we found one instance (ASC9) where an increase in LGA (17 vs 13) placed an individual’s sequential sample among the highest genome instability scores, along with the HRm samples. However, the heightened LGA did not cross the threshold of 20 described by Eeckhoutte *et al* (2020) to indicate HRD ^32^.

#### 1.1.1 Opportunistic liquid biopsy versus excisional tissue biopsy

Among our cohort, 3 participants had samples collected at 3 timepoints (2xcfDNA and 1x FFPE), each with >800 days between the first and last sampling. We observed differences in cancer cell frequency of clones identified in these samples with different time points and collection approaches (Figure 4A). As artefactual variants were reported in ASC28FFPE, the validity of the uniquely reported clones in that sample is unclear.

**Figure 4.**
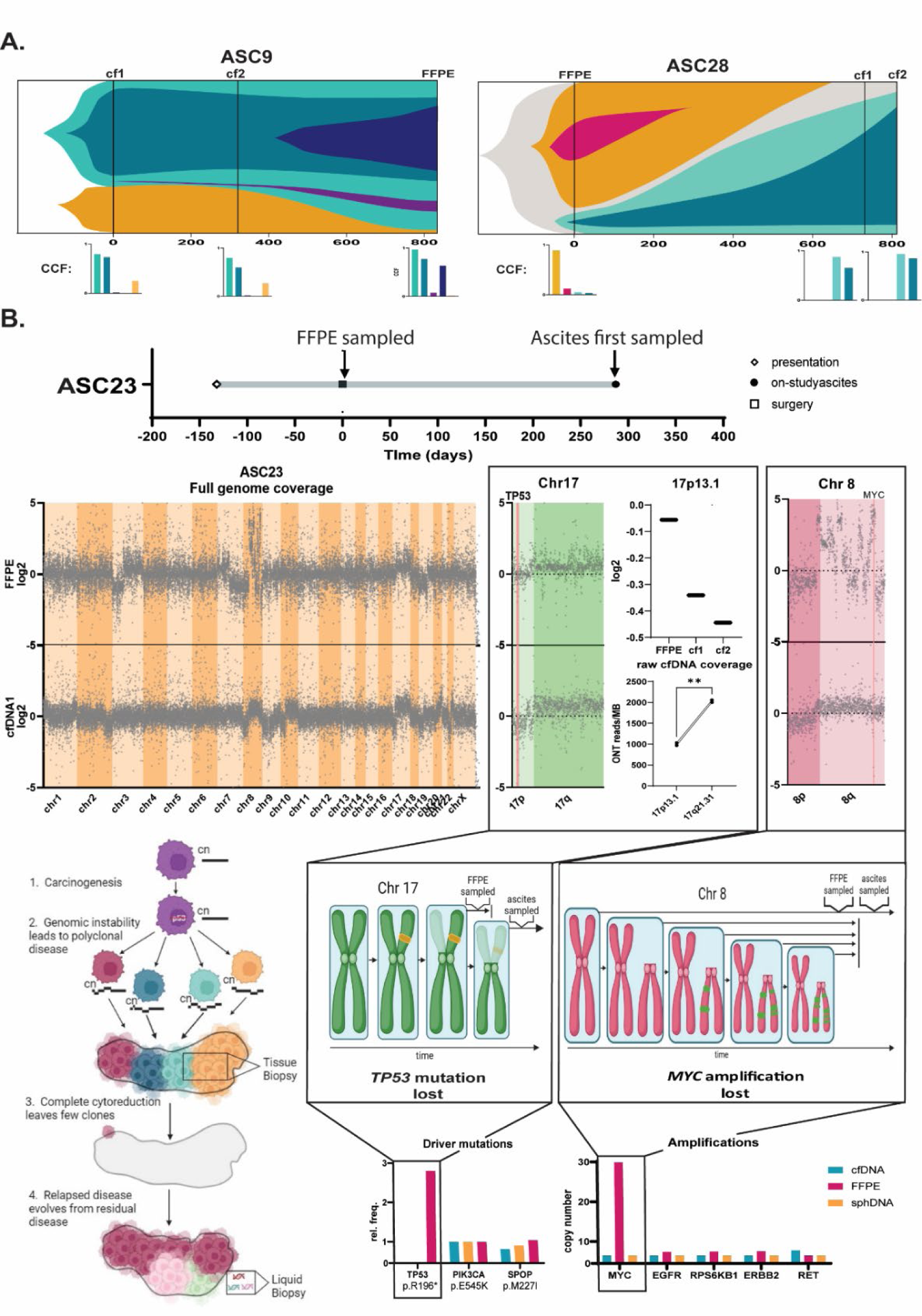
Differences over time in liquid and tissue biopsies. (A) Representations of estimated clonal evolution in 2 samples based on cluster cancer cell frequencies indicated below at each of three timepoints. (B) Log2 read depth in cfDNA1 and FFPE samples collected from participant ASC23 nearly 10 months apart with higher resolution reproductions of Chr17 and Chr8. Disparities in Chr17 are additionally represented by log2 of all variants on Chr17p13.1 in FFPE and serial ascites samples and a deletion of 17p13.1 is verified by paired t-test of ONT reads per MB on 17p13.1 and 17q21.31 in serial cfDNA samples (raw cfDNA coverage, p=0.003). Mechanisms of these temporal changes are modelled above the clinical reporting consequences (and reporting references). cfDNA, cell-free DNA, FFPE, formalin-fixed paraffin embedded; CCF, cancer cell fraction; ONT, Oxford Nanopore Technology Sequencing.

Though we were unable to reliably track clones in participant ASC23, due to a larger number of confounding artefactual variants in the FFPE, we did note some key differences between samples (Figure 4B). Specifically, CN profiling revealed discordance between FFPE and ascites samples, explaining previously noted differences in *TP53* mutation and *MYC* amplification (Figure 2A). Notably, chr17p13.1 features a low log2 (-0.056) in the FFPE (consistent with the double-hit hypothesis ^36^), but significantly lower log2 (-0.34 and -0.44) in the ascites samples collected approximately 10 and 28 months later, respectively, suggesting a clonal loss of the second 17p13.1, along with the *TP53* mutation. This was confirmed with a decrease of alignment to 17p13.1 in cfDNA samples, relative to similarly sized and covered 17q21.31, by native ONT whole genome sequencing (paired t-test, p=0.003). Chromosome 8q demonstrates severe instability in the FFPE sample, including a MYC amplification of 13x (CN >29), indicative of chromothripsis, which is not maintained in ascites samples. Theses disparities suggest that the clones captured in the FFPE were less prevalent when ascites was collected than would be indicated by analysing the archived FFPE biopsy, likely due to a combination of tumour evolution and sampling bias (Figure 4B).

## 4. Discussion

cfDNA in ascites offers a clear opportunity for representative and not-additionally invasive profiling of the ovarian cancer genome, enabling facilitation of precision medicine where tissue biopsies are contraindicated or compromised. This study has examined the feasibility, reliability and reproducibility of targeted sequencing output from ascites cfDNA and has captured unique evidence of tumour evolution identifiable with longitudinal sampling.

This study demonstrates that quality, representative sequencing data can be attained from ascites cfDNA. cfDNA was observed to be high in tumour content (up to 95%) with mutation profiles concordant with solid tumours, demonstrated by comparison to archived FFPE tissue, clinical reports and the Cancer Genome Atlas ^34^. Mutations of key clinical relevance included 3 somatic *BRCA1/2* cases identifiable in cfDNA. Importantly, HRD was assessable in cfDNA. HRD samples with sufficient tumour content could be identified by LOH, LGAs and high TMB, extending on methods previously used for HRD assessment in cfDNA ^16, 17^. Kfoury *et al.* (2023) and Roussel-Simonin *et al.* (2023) recently reported a strong correlation in HRD scores between cfDNA and tissue biopsies ^16, 17^.

Among our cohort, ascites sampling preceded surgery in 7 of 15 participants, in one case by over 2 years. This was particularly key in 2 of the 3 cases with somatic *BRCA1/2* mutations, where ascites preceded surgery by 100 days or more. As sequencing turnaround time can be substantial ^16^, earlier knowledge of *BRCA1/2* mutations and HRD may be instrumental in the streamline delivery of maintenance PARPi or application of neoadjuvant PARPi, which is currently under investigation for efficacy ^37^. Core biopsies have a >25% failure rate for provision of adequate quality sequencing template ^38^, so ascites cfDNA may offer the most robust and timely alternative in the frontline setting.

In 9 of the 15 cases studied (to date), ascites emerged with disease relapse, where clinical trials of targeted therapies are often considered and archived tissue is accessed for biomarker profiling ^39^. We have demonstrated the value of opportunistic ascites sampling by showing evidence of tumour evolution identifiable in sequential samples. Genome instability markers and CN profiles were seen to change over time, in line with previous reports ^24^. Furthermore, >10% shifts in clonal prevalence were observed in 5 of 10 sequential ascites sample pairs. This, along with our key finding of a high prevalence of sequencing artefacts in 2 of 5 archived FFPE samples, demonstrates the improved likelihood of identifying and ascertaining the relevance of clinically actionable variants in ascites over archived FFPE.

One case showed particularly prevalent divergence between samples. When comparing FFPE from primary debulking surgery and cfDNA at disease relapse, deletion of both Chr17p13.1 and a chromothripsis-affected Chr8q was observed, along with other significant CN changes. These changes suggest active clonal selection throughout the course of the disease. As well as evolution, the limitation of sampling bias that is inherent to tissue biopsies may have played a part ^40^. Ascites cfDNA, which flows freely through the peritoneal cavity, and is sampled in an unbiased manner, overcomes this limitation ^40^. cfDNA is likely proportionally representative of peritoneal lesions, capturing intra-tumour heterogeneity of ovarian cancer, which is usually peritoneally confined ^41^.

A strength of the present study is our report of the technical performance of cfDNA, using a commercially available NGS platform, not optimised for use with cfDNA. cfDNA achieved significantly improved coverage over sphDNA and FFPE, subsequently improving capacity for variant identification and verification. Additionally, when we performed CN profiling on sequential cfDNA samples across the cohort, 10 of 11 sample pairs clustered within participants, demonstrating reproducibility, with a trend towards more divergence in samples with extensive intervening chemotherapy. sphDNA similarly clustered in participant groups, however 3 of 5 FFPE samples incorrectly clustered, potentially indicative of susceptibility to CN estimation errors. In the case study we performed of duplicate cfDNA sequencing, consensus in variant detection and CN profile was observed.

The main limitation of this work was the small cohort size, hampering our ability to observe statistically significant differences. A larger cohort, covering a broader range of ovarian cancer histological subtypes, would provide greater confidence in our findings. We were also unable to compare ascites with fresh tissue as in most cases ascites was collected at routine paracentesis. This limited our assessment of concordance with tumour tissue, however, published works have previously demonstrated high similarity between ascites cfDNA and time-matched fresh tumour tissue ^15-17^. As ascites is often a contraindication for surgery and the accompanying opportunity for excisional biopsy, we believe the comparison with archived tissue is a better imitation of the clinical setting. Additionally, although this work demonstrated successful application of several bioinformatic analyses often reserved for whole genome-spanning data sets, we were unable to compute clinically-analogous HRD scores in the absence of either whole genome sequencing data or matched normal tissue DNA.

Future studies with larger cohorts and more samples for spatial, temporal and genomic reference could provide further confidence in cfDNA analysis, particularly for HRD assessment. A recent report showcased the high spatial and temporal variability of HRD scores in high grade serous ovarian cancer samples ^42^. Considering the unbiased sampling possible with cfDNA analysis, it should be investigated whether cfDNA-based HRD analysis can encapsulate the most reflective HR status at key time points and across tumour sites, allowing better prediction of PARPi efficacy. Similar assessment should be performed on new molecular biomarkers for targeted treatments, as they continue to emerge.

This study demonstrates the potential benefit for opportunistic sequencing of cfDNA from ascites to guide personalised medicine for ovarian cancer. We show cfDNA to provide exemplary template for targeted sequencing; being high in tumour content, producing no sequencing artefact and identifying key disease markers which evolve over time. With these findings supporting several recent publications, we argue for the immediate implementation of ascites cfDNA sequencing to inform precision medicine in lieu of invasive tissue biopsies.

## Conclusion

This research indicates the suitability of cfDNA from ovarian cancer ascites for somatic mutation profiling and genome instability inference, to facilitate precision medicine. We show cfDNA to reliably identify clinically actionable information at key timepoints, using an opportunistic, not-additionally invasive surrogate biopsy approach. This is of particular relevance at initial diagnosis where ascites precedes interval debulking surgery, or at disease relapse as an alternative to profiling archived formalin-fixed tissue. The evidence presented here aligns with various recent reports, highlighting the utility of ascites fluid for genomic profiling in the clinical setting.

## Data Availability

All deidentified data produced in the present study may be made available upon reasonable request to the authors.

## Acknowledgements

We would firstly like to acknowledge the participants of this study and their families for their involvement. We would also like to acknowledge our consumer partners, Gill Stannard, Kathryn Leaney, Gillian Begbie and Jacinta Frawley, for their insight and support of this study. We are also grateful to Caroline Maria Rossing for providing critical evaluation of a draft of this manuscript. The research presented was supported by a number of anonymous philanthropic donations, and in part by the Intramural Research Program of the NIH and the Adam J Berry Memorial Fund through the Australian Academy of Science. BW is supported by a research training program scholarship from the Australian Government and a PhD top-up scholarship from SPHERE Cancer CAG, supported by Cancer Institute NSW. We acknowledge contributions made to this research by the Health Systems Alliance Biobank for provision of some samples. Some components of Figures 1 and 4 were generated using Biorender.com.

## Author Contributions

Conceptualisation, B.W., K.W., C.E.F, Formal analysis, B.W., M.D., K.W., C.E.F., Investigation, B.W., E.P., Resources, J.D., Y.C.L., V.A., R.A.,M.D., Data Curation, B.W., M.C., M.D., Writing – original draft, B.W., Writing – review and editing, B.W., E.P., J.D., M.C.,Y.C.L., V.A.,R.A.,M.D.,K.W.,C.E.F., Visualisation, B.W., Supervision, M.C, M.D., K.W., C.E.F, Funding acquisition, C.E.F., M.D.

## References

1. Foo T, George A, Banerjee S. PARP inhibitors in ovarian cancer: An overview of the practice-changing trials. Genes Chromosomes Cancer. 2021;60(5):385–97.

2. Gonzalez-Martin A, Harter P, Leary A, Lorusso D, Miller RE, Pothuri B, Ray-Coquard I, Tan DSP, Bellet E, Oaknin A, et al. Newly diagnosed and relapsed epithelial ovarian cancer: ESMO Clinical Practice Guideline for diagnosis, treatment and follow-up. Ann Oncol. 2023;34(10):833–48.

3. Coleman RL, Spirtos NM, Enserro D, Herzog TJ, Sabbatini P, Armstrong DK, Kim JW, Park SY, Kim BG, Nam JH, et al. Secondary Surgical Cytoreduction for Recurrent Ovarian Cancer. N Engl J Med. 2019;381(20):1929–39.

4. Hoang LN, Zachara S, Soma A, Kobel M, Lee CH, McAlpine JN, Huntsman D, Thomson T, van Niekerk D, Singh N, et al. Diagnosis of Ovarian Carcinoma Histotype Based on Limited Sampling: A Prospective Study Comparing Cytology, Frozen Section, and Core Biopsies to Full Pathologic Examination. Int J Gynecol Pathol. 2015;34(6):517–27.

5. Huang H, Li YJ, Lan CY, Huang QD, Feng YL, Huang YW, Liu JH. Clinical significance of ascites in epithelial ovarian cancer. Neoplasma. 2013;60(5):546–52.

6. Anuradha S, Webb PM, Blomfield P, Brand AH, Friedlander M, Leung Y, Obermair A, Oehler MK, Quinn M, Steer C, et al. Survival of Australian women with invasive epithelial ovarian cancer: a population-based study. Med J Aust. 2014;201(5):283–8.

7. Quan Q, Zhou S, Liu Y, Yin W, Liao Q, Ren S, Zhang F, Meng Y, Mu X. Relationship between ascites volume and clinical outcomes in epithelial ovarian cancer. J Obstet Gynaecol Res. 2021.

8. Krugmann J, Schwarz CL, Melcher B, Sterlacci W, Vieth M, Rosch S, Lermann J. Diagnostic impact of ascites cytology in 941 patients: malignancy rates and time of detection in ovarian cancer relative to other tumor types. Arch Gynecol Obstet. 2020;301(6):1521–32.

9. Baransi S, Michaan N, Gortzak-Uzan L, Aizic A, Laskov I, Gamzu R, Grisaru D. The accuracy of ascites cytology in diagnosis of advanced ovarian cancer in postmenopausal women prior to neoadjuvant chemotherapy. Menopause. 2020;27(7):771–5.

10. Swisher EM, Wollan M, Mahtani SM, Willner JB, Garcia R, Goff BA, King MC. Tumor-specific p53 sequences in blood and peritoneal fluid of women with epithelial ovarian cancer. Am J Obstet Gynecol. 2005;193(3 Pt 1):662-7.

11. Tadic V, Zhang W, Brozovic A. The high-grade serous ovarian cancer metastasis and chemoresistance in 3D models. Biochim Biophys Acta Rev Cancer. 2024;1879(1):189052.

12. Werner B, Yuwono N, Duggan J, Liu D, David C, Srirangan S, Provan P, Investigators IN, DeFazio A, Arora V, et al. Cell-free DNA is abundant in ascites and represents a liquid biopsy of ovarian cancer. Gynecol Oncol. 2021;162(3):720–7.

13. Allan Z, Witts S, Tie J, Tebbutt N, Clemons NJ, Liu DS. The prognostic impact of peritoneal tumour DNA in gastrointestinal and gynaecological malignancies: a systematic review. Br J Cancer. 2023.

14. Ibanez de Caceres I, Battagli C, Esteller M, Herman JG, Dulaimi E, Edelson MI, Bergman C, Ehya H, Eisenberg BL, Cairns P. Tumor cell-specific BRCA1 and RASSF1A hypermethylation in serum, plasma, and peritoneal fluid from ovarian cancer patients. Cancer Res. 2004;64(18):6476–81.

15. Han MR, Lee SH, Park JY, Hong H, Ho JY, Hur SY, Choi YJ. Clinical Implications of Circulating Tumor DNA from Ascites and Serial Plasma in Ovarian Cancer. Cancer Res Treat. 2020;52(3):779–88.

16. Roussel-Simonin C, Blanc-Durand F, Tang R, Vasseur D, Le Formal A, Chardin L, Yaniz E, Gouy S, Maulard A, Scherier S, et al. Homologous recombination deficiency (HRD) testing on cell-free tumor DNA from peritoneal fluid. Mol Cancer. 2023;22(1):178.

17. Kfoury M, Hazzaz RE, Sanson C, Durand FB, Michels J, Blameble EC, Tang R, Le Formal A, Lecerf E, Gouy S, et al. Circulating Tumor DNA from Ascites as an alternative to tumor sampling for genomic profiling in ovarian cancer patients. Biomark Res. 2023;11(1):93.

18. du Bois A, Reuss A, Pujade-Lauraine E, Harter P, Ray-Coquard I, Pfisterer J. Role of surgical outcome as prognostic factor in advanced epithelial ovarian cancer: a combined exploratory analysis of 3 prospectively randomized phase 3 multicenter trials: by the Arbeitsgemeinschaft Gynaekologische Onkologie Studiengruppe Ovarialkarzinom (AGO-OVAR) and the Groupe d’Investigateurs Nationaux Pour les Etudes des Cancers de l’Ovaire (GINECO). Cancer. 2009;115(6):1234–44.

19. Nasioudis D, Byrne M, Ko EM, Haggerty AF, Cory L, Giuntoli Ii RL, Kim SH, Latif NA. Ascites volume at the time of primary debulking and overall survival of patients with advanced epithelial ovarian cancer. Int J Gynecol Cancer. 2021;31(12):1579–83.

20. Vergote I, Trope CG, Amant F, Kristensen GB, Ehlen T, Johnson N, Verheijen RH, van der Burg ME, Lacave AJ, Panici PB, et al. Neoadjuvant chemotherapy or primary surgery in stage IIIC or IV ovarian cancer. N Engl J Med. 2010;363(10):943–53.

21. Corcoran RB, Chabner BA. Application of Cell-free DNA Analysis to Cancer Treatment. N Engl J Med. 2018;379(18):1754–65.

22. Al-Kateb H, Nguyen TT, Steger-May K, Pfeifer JD. Identification of major factors associated with failed clinical molecular oncology testing performed by next generation sequencing (NGS). Mol Oncol. 2015;9(9):1737–43.

23. Zalaznick H. CB, Cogan E., Perry M., Trost J., Mancini-DiNardo D., Gutin A., Lanchbury J., Timms K. Rates of homologous recombination deficiency across different subtypes of ovarian cancer and in pre- and post-neoadjuvant chemotherapy tumor samples (139.5). Gyneocologic Oncology. 2022;166:S86–S7.

24. Burdett NL, Willis MO, Alsop K, Hunt AL, Pandey A, Hamilton PT, Abulez T, Liu X, Hoang T, Craig S, et al. Multiomic analysis of homologous recombination-deficient end-stage high-grade serous ovarian cancer. Nat Genet. 2023;55(3):437–50.

25. Patch AM, Christie EL, Etemadmoghadam D, Garsed DW, George J, Fereday S, Nones K, Cowin P, Alsop K, Bailey PJ, et al. Whole-genome characterization of chemoresistant ovarian cancer. Nature. 2015;521(7553):489-94.

26. Lau JW, Lehnert E, Sethi A, Malhotra R, Kaushik G, Onder Z, Groves-Kirkby N, Mihajlovic A, DiGiovanna J, Srdic M, et al. The Cancer Genomics Cloud: Collaborative, Reproducible, and Democratized-A New Paradigm in Large-Scale Computational Research. Cancer Res. 2017;77(21):e3–e6.

27. Church DM, Schneider VA, Graves T, Auger K, Cunningham F, Bouk N, Chen HC, Agarwala R, McLaren WM, Ritchie GR, et al. Modernizing reference genome assemblies. PLoS Biol. 2011;9(7):e1001091.

28. Tamborero D, Rubio-Perez C, Deu-Pons J, Schroeder MP, Vivancos A, Rovira A, Tusquets I, Albanell J, Rodon J, Tabernero J, et al. Cancer Genome Interpreter annotates the biological and clinical relevance of tumor alterations. Genome Med. 2018;10(1):25.

29. Oscanoa J, Sivapalan L, Gadaleta E, Dayem Ullah AZ, Lemoine NR, Chelala C. SNPnexus: a web server for functional annotation of human genome sequence variation (2020 update). Nucleic Acids Res. 2020;48(W1):W185–W92.

30. Infrastructure) PR. Katana. UNSW Sydney2010.

31. Talevich E, Shain AH, Botton T, Bastian BC. CNVkit: Genome-Wide Copy Number Detection and Visualization from Targeted DNA Sequencing. PLoS Comput Biol. 2016;12(4):e1004873.

32. Eeckhoutte A, Houy A, Manie E, Reverdy M, Bieche I, Marangoni E, Goundiam O, Vincent-Salomon A, Stoppa-Lyonnet D, Bidard FC, et al. ShallowHRD: detection of homologous recombination deficiency from shallow whole genome sequencing. Bioinformatics. 2020;36(12):3888–9.

33. Gillis S, Roth A. PyClone-VI: scalable inference of clonal population structures using whole genome data. BMC Bioinformatics. 2020;21(1):571.

34. Cancer Genome Atlas Research N. Integrated genomic analyses of ovarian carcinoma. Nature. 2011;474(7353):609-15.

35. Coleman RL, Oza AM, Lorusso D, Aghajanian C, Oaknin A, Dean A, Colombo N, Weberpals JI, Clamp A, Scambia G, et al. Rucaparib maintenance treatment for recurrent ovarian carcinoma after response to platinum therapy (ARIEL3): a randomised, double-blind, placebo-controlled, phase 3 trial. Lancet. 2017;390(10106):1949–61.

36. Liu Y, Chen C, Xu Z, Scuoppo C, Rillahan CD, Gao J, Spitzer B, Bosbach B, Kastenhuber ER, Baslan T, et al. Deletions linked to TP53 loss drive cancer through p53-independent mechanisms. Nature. 2016;531(7595):471-5.

37. Westin S, Michael V, Fellman B, Meyer L, Taylor T, Alvarado T, Grinsfelder M, Fleming N, Shafer A, Cobb L, et al. Now: Neoadjuvant Olaparib window trial in patients with newly diagnosed BRCA mutant ovarian cancer. Gynecologic Oncology. 2023;176:S2–S3.

38. Turashvili G, Lazaro C, Ying S, Charames G, Wong A, Hamilton K, Yee D, Agro E, Chang M, Pollett A, et al. Tumor BRCA Testing in High Grade Serous Carcinoma: Mutation Rates and Optimal Tissue Requirements. Cancers (Basel). 2020;12(11).

39. Ethier JL, Fuh KC, Arend R, Konecny GE, Konstantinopoulos PA, Odunsi K, Swisher EM, Kohn EC, Zamarin D. State of the Biomarker Science in Ovarian Cancer: A National Cancer Institute Clinical Trials Planning Meeting Report. JCO Precis Oncol. 2022;6:e2200355.

40. Paracchini L, D’Incalci M, Marchini S. Liquid Biopsy in the Clinical Management of High-Grade Serous Epithelial Ovarian Cancer-Current Use and Future Opportunities. Cancers (Basel). 2021;13(10).

41. Thomakos N, Diakosavvas M, Machairiotis N, Fasoulakis Z, Zarogoulidis P, Rodolakis A. Rare Distant Metastatic Disease of Ovarian and Peritoneal Carcinomatosis: A Review of the Literature. Cancers (Basel). 2019;11(8).

42. Cunnea P, Curry EW, Christie EL, Nixon K, Kwok CH, Pandey A, Wulandari R, Thol K, Ploski J, Morera-Albert C, et al. Spatial and temporal intra-tumoral heterogeneity in advanced HGSOC: Implications for surgical and clinical outcomes. Cell Rep Med. 2023;4(6):101055.

