## Supplementary File 1 for "Cell-free DNA from ascites identifies clinically relevant variants and tumour evolution in patients with advanced ovarian cancer"

**Supplementary File 1. Extended methods**

Data processing

TSO500 sequencing output was processed using the TSO500 v2.2 Local Application. Briefly, binary base calls were converted into FASTQ files and aligned to GRCh37, with UMI read collapsing, using the Burrows-Wheeler Aligner with the SAM Tools utility. One FFPE sample achieved MEC below the guideline of 150x (ASC17FFPE, MEC 105). The data from the 300-cycle run was trimmed to be 2x100 bp for analysis. Small variants (single nucleotide polymorphisms (SNPs), multi-nucleotide polymorphisms (MNPs) and insertions/deletions (indels) were called using Pisces software and filtered using Pepe software. Filtered variants, which passed quality assessment, were annotated by the Illumina Annotation Engine Nirvana with reference to databases including dbSNP, gnomAD genome and exome, 1000 genomes, ClinVar, COSMIC, RefSeq, and Ensembl. Gene amplifications were investigated only in specific genes, as listed in TSO500 literature, by CRAFT copy number variant calling software.

Somatic variant verification and cancer driving classification

Total variants were counted and samples with total variant count outside of a Z-score (from a mean excluding one majorly outlying sample) of |3| were identified. Variants were considered verifiable where reported in >1 sample per participant, or if unique, where >0.05 variant allele frequency (VAF) and >150x read depth.

We focussed our analysis on non-synonymous variants with a minor allele count of less than 100 in each of three population databases (<0.05% GnomAD Exome, <0.5% GnomAD Genome and <2% 1000 Genomes). As we had no germline references to compare to, these variants were considered ‘potentially somatic’ (unless known to be germline based on clinical reports) and were used for mutational signature assessment.

SNPs and MNPs which met these criteria were assessed by the Cancer Genome Interpreter (CGI) [1], using SNPnexus [2], to identify their likelihood of a cancer driving capacity. Insertions, deletions or SNPs with no annotation on CGI were searched for likelihood of pathogenicity using ClinVar, Varsome and COSMIC.

For people with pathogenic variants in HRD-related genes, reversion mutations were searched for manually. Any variant reported in driver HRD gene with low population and/or allele frequency was investigated on Integrated Genome Viewer (IGV). Variants which restored reading frame, corrected premature stop codon or restored a missense codon to synonymous were considered potentially reversion mutations, particularly if on the same read as pathogenic variant. No likely reversion mutations were identified, we found only a low frequency (max. 0.02VAF) additional BRCA1 synonymous SNV downstream of nonsense mutation in ASC20 and a high frequency (0.87 VAF) BRCA1 missense SNV ASC38 which did not restore reading frame or premature stop codon.

Tumour purity

Tumour purity of samples was estimated based on the typical trajectory of HGSOC carcinogenesis, where if a single deleterious *TP53* mutation was present, the competing healthy allele is deleted conforming to the double-hit hypothesis [3]. Thus, VAF of a deleterious *TP53* mutation was considered to represent 1n cancer cells, and these were considered as a proportion of 1n cancer cells + 2n healthy cells, by the following formula (where *t* is tumour fraction and *v* is the VAF of the deleterious *TP53* variant).

$$t=\frac{1-v}{2}$$

For patient ASC23, tumour purity was estimated based on proportionate deleterious *PIK3CA* variant VAF in cfDNA and sphDNA compared to FFPE, made relative to FFPE estimated tumour purity (based on *TP53*). This sample was excluded from clonal analysis to avoid errors from miscalculated purity.

Mutational signature analysis

All ‘potentially somatic’ variants with read depth > 150x (aside from sample ASC17FFPE, where the average readdepth was below 150, so all potentially somatic variants were tested) were profiled by SigProfiler tools, SigProfiler Matrix Generator and SigProfiler Extractor, using the computational cluster Katana [4].

Tumour mutational burden (TMB)

The TMB of each sample was determined by the TMB Trace Function of the TSO-500 v2.2 Local App. This function includes only variants with a high confidence of being somatic, dividing the number of eligible variants by the effective panel size (as described in the TSO-500 v2.2 Local App User Guide).

Gene-specific copy number variants

Gene amplifications, reported as fold-change by the TSO500 Local App were used to estimate copy number by the following formula (where $n$ is the copy number, $x$ is the reported fold-change and $y$ is the estimated tumour purity):

$$n=\frac{200x-2\left( 100-y \right)}{y}$$

Copy number variants

To estimate variance from a normal diploid state we used CNVKit [5], a program designed to calculate copy number from targeted sequencing data, using the computational cluster Katana [4]. CNVKit was calibrated with the TSO500 probe BED file, identifying 30624 sites with high on- and off-target coverage when using the TSO500 sequencing panel. For each sample, the log2 copy ratio of read depth at each site was calculated and used to estimate local copy number. Profiles were assessed against a flat reference and output was rescaled based on estimated tumour purity. Comparison of log2 ratios across sites were used to identify unique copy number segments, identifying breakpoints for amplifications, deletions and structural changes [6]. The number of unique segments identified were counted (including only segments where read depth was within a modified Z-score of |3.5|). To compare genome-wide segmental copy number between samples and individuals, we used the ‘Clustermap’ function of Seaborn Python package to cluster and compute the pair-wise distance between samples . For this genome-wide analysis, where sites were within segments exceeding the described modified z-score, these sites were assigned the average of the neighbouring segments’ estimated copy number.

Large-scale genome alterations (LGA)

From unique segments identified, segments were counted as large-scale genome alterations if they were >10MB in length and adjacent to other segments >10MB on the same chromosome. LGA counts ≥20 were indicative of high genome instability, counts 15-20 were borderline and counts ≤14 were low instability [7].

Loss of heterozygosity

Loss of heterozygosity (LOH) was identified by a deviation of average variant allele frequency from an expected range of 0.4-0.6VAF amongst non-homozygous small variants, with read depth >150x (0.05<VAF<0.95), within chromosome-limited bins of 100KB, as a percentage of all bins where variants were located. Where average read depth was below 150x (ASC017FFPE, ASC030FFPE) or where median VAF was below 0.1, LOH% could not reliably be determined (ASC023FFPE).

Genomic instability

LGAs, LOH and TMB were considered indicators of genomic stability. The median values of each indicator (among initial cfDNA samples) were considered the threshold and individuals received a score (0-3) counting the number of indicators in which their values were above the threshold.

Clonal analysis – cluster identification

PyClone-VI was used to cluster somatic variants into clone groups and estimate the cancer cell frequency of the clone groups in each sample of a participant’s set [8]. VAF, read depth, estimated copy number, estimated tumour purity and expected normal copy (diploid) of potentially somatic variants were used as input. Beta-binomial distribution and 10 random restarts were used. Variants with copy number of 0 in any sample could not be included in analysis. In one case (ASC28) this included the *TP53* variant, so a high frequency unrepresented clone was assumed, coloured grey. Identified clusters in which VAF of all included variants was stable around 0.5 in all samples were assumed to include misidentified heterozygous germline variants and were excluded.

To limit clonal analysis to high confidence somatic mutations, where participants had any samples of <30% tumour content, only variants of below 0.3 VAF in that sample were included in clonal analysis. Variants which were not reported across all samples were screened on IGV to identify read depth and VAF.

Clonal analysis – cluster prevalence evolution

Cluster phylogeny and evolution was estimated logically based on the relationships between CCFs across timepoints. Changes in CCF were reported when >10% shifts seen between sequential cfDNA samples.

[1]	Tamborero, D. *et al.* Cancer Genome Interpreter annotates the biological and clinical relevance of tumor alterations. *Genome Med* **10**, 25, doi:10.1186/s13073-018-0531-8 (2018).

[2]	Oscanoa, J. *et al.* SNPnexus: a web server for functional annotation of human genome sequence variation (2020 update). *Nucleic Acids Res* **48**, W185-W192, doi:10.1093/nar/gkaa420 (2020).

[3]	Liu, Y. *et al.* Deletions linked to TP53 loss drive cancer through p53-independent mechanisms. *Nature* **531**, 471-475, doi:10.1038/nature17157 (2016).

[4]	Katana (UNSW Sydney, 2010).

[5]	Talevich, E., Shain, A. H., Botton, T. & Bastian, B. C. CNVkit: Genome-Wide Copy Number Detection and Visualization from Targeted DNA Sequencing. *PLoS Comput Biol* **12**, e1004873, doi:10.1371/journal.pcbi.1004873 (2016).

[6]	Venkatraman, E. S. & Olshen, A. B. A faster circular binary segmentation algorithm for the analysis of array CGH data. *Bioinformatics* **23**, 657-663, doi:10.1093/bioinformatics/btl646 (2007).

[7]	Eeckhoutte, A. *et al.* ShallowHRD: detection of homologous recombination deficiency from shallow whole genome sequencing. *Bioinformatics* **36**, 3888-3889, doi:10.1093/bioinformatics/btaa261 (2020).

[8]	Gillis, S. & Roth, A. PyClone-VI: scalable inference of clonal population structures using whole genome data. *BMC Bioinformatics* **21**, 571, doi:10.1186/s12859-020-03919-2 (2020).
