## Supplementary Figures & Tables for "Cell-free DNA from ascites identifies clinically relevant variants and tumour evolution in patients with advanced ovarian cancer"

**Supplementary Table 1. Cohort details**

| **Research ID** | **Histotype** | **Chemotherapy history at recruitment** |  | **Tissue sequenced** | **Days since first cfDNA** | **Chemotherapy between cfDNA samples** |
| --- | --- | --- | --- | --- | --- | --- |
| *ASC6* | HGS | Naïve |  | cfDNA (1) | n/a | n/a |
| *ASC7* | HGS | Carboplatin |  | ser. cfDNA (2) | 265 | Paclitaxel, gemcitibine |
| *ASC9* | HGS | Naïve |  | ser. cfDNA (2)  sphDNA (1)  FFPE (1) | 319  0  831 | Carboplatin, paclitaxel, pegylated liposomal doxorubicin, bevacizumab |
| *ASC10* | HGS | Carboplatin, pegylated liposomal doxorubicin, tamoxifen |  | ser. cfDNA (2) | 22 | Nil |
| *ASC11* | HGS | Carboplatin, paclitaxel, veliparib/placebo, tamoxifen, pegylated liposomal doxorubicin, olaparib |  | ser. cfDNA (2)  sphDNA (1) | 36  0 | Carboplatin, pegylated liposomal doxorubicin, paclitaxel |
| *ASC12* | HGS | Naïve |  | ser. cfDNA (2) | 13 | Carboplatin, paclitaxel, durvalumab, tremelimumab |
| *ASC17* | HGS | Naïve |  | cfDNA (1)  FFPE (1) | n/a  103 | n/a |
| *ASC20* | HGS | Carboplatin, paclitaxel, cisplatin, pegylated liposomal doxorubicin, olaparib, trabectidin |  | ser. cfDNA (2)  sphDNA (1) | 28  0 | Cyclophosphamide |
| *ASC23* | Clear cell | Naïve |  | ser. cfDNA (2)  sphDNA (1)  FFPE (1) | 559  0  -287 | Carboplatin, paclitaxel, bevacizumab, THOR-707, pegylated liposomal doxorubicin, bevacizumab, olaparib, CYH33 (PIK3k inhibitor) |
| *ASC26* | HGS | Naïve |  | ser. cfDNA (2) | 21 | Carboplatin, paclitaxel |
| *ASC28* | HGS | Carboplatin, paclitaxel, bevacizumab, pegylated liposomal doxorubicin |  | ser. cfDNA (2)  sphDNA (1)  FFPE (1) | 78  0  -732 | Nil |
| *ASC30* | HGS | Carboplatin, paclitaxel, bevacizumab |  | dup. cfDNA (2)  FFPE (1) | 0  0 | n/a |
| *ASC36* | HGS | Naïve |  | ser. cfDNA (2) | 17 | Carboplatin, paclitaxel |
| *ASC37* | HGS | Carboplatin, paclitaxel, pegylated liposomal doxorubicin, olaparib cediranib/placebo, doxorubicin, bevacizumab |  | ser. cfDNA (2) | 99 | Carboplatin, gemcitabine |
| *ASC38* | HGS | Naive |  | cfDNA (1) | n/a | n/a |

*HGS, high grade serous; cfDNA, ascites cell-free DNA; sphDNA, ascites spheroid cell DNA; FFPE, formalin-fixed paraffin embedded tissue section DNA; ser., serial; dup., duplicate; n/a, not applicable;*

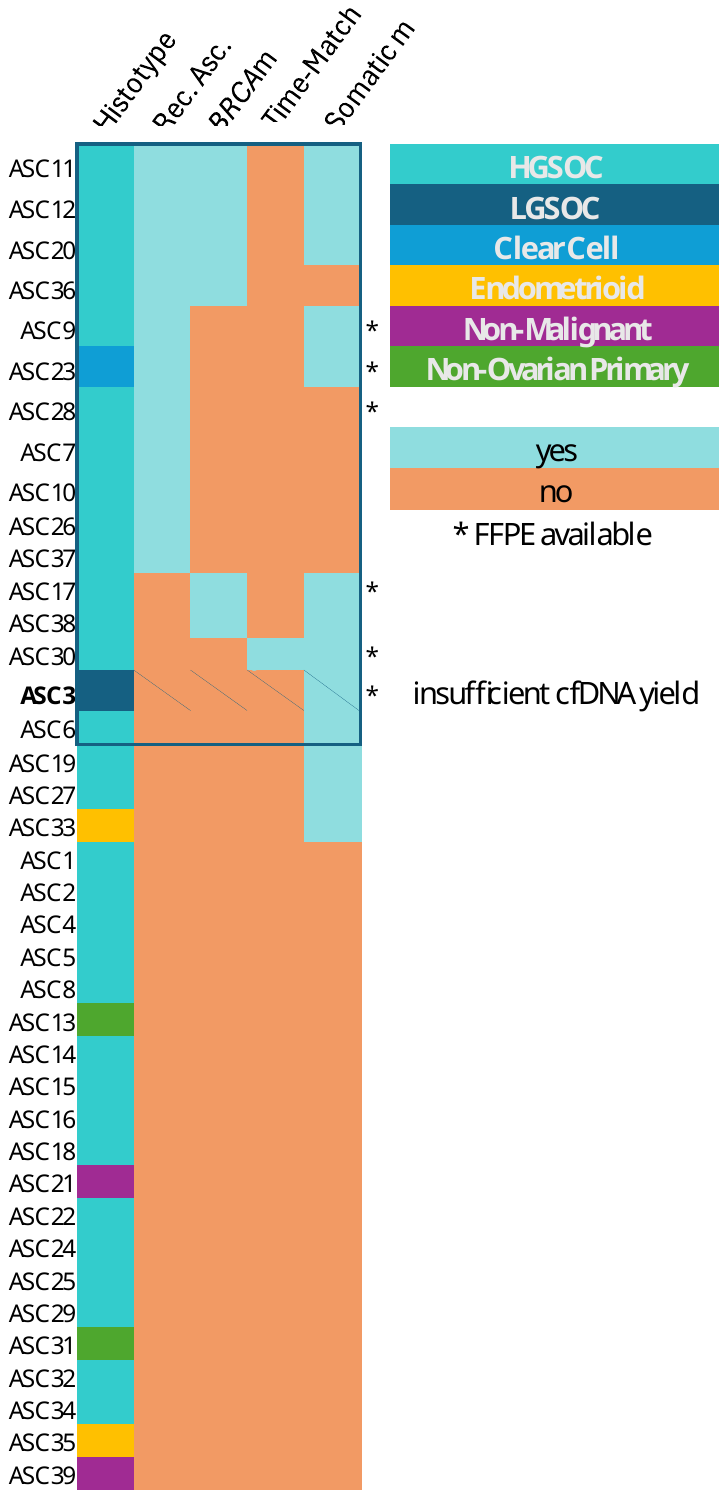

**Supplementary Figure 1.** *The cohort was selected based on meeting any of these criteria in order of priority: recurrent ascites (Rec. Asc.), i.e. ascites collected at multiple timepoints, BRCA mutations in clinical report (BRCAm), time matched tissue sample from surgery available, non-BRCA somatic mutations in clinical report. The first 15 people to meet these criteria and have sufficient sample were included.*

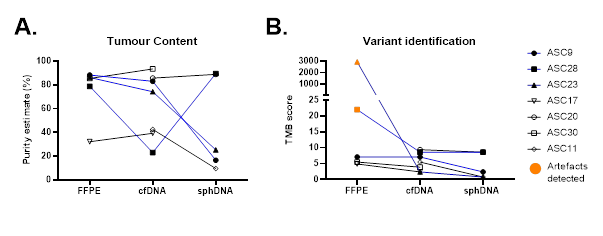

**Supplementary Figure 2.** *Estimated tumour purity (A) and TMB score (B) in cfDNA compared to sample-matched sphDNA and non-time-matched FFPE. Participants with full set available are identified by filled shapes and blue lines. Orange markers (B) indicate samples where artefactual single base substitution signatures were identified. cfDNA, cell-free DNA; sphDNA, DNA from ascites-derived cell spheroids; FFPE, DNA from formaldehyde-fixed paraffin embedded tumour biopsy samples.*

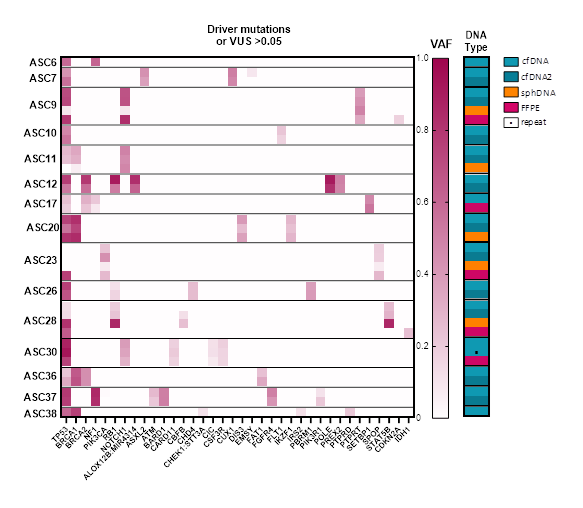
**Supplementary Figure 3.** *Variant allele frequency and consensus between samples in cancer driving mutations with >5% VAF.*

**Supplementary Table 2. Allele Frequency of clinically reported variants**

| ***Patient ID*** | ***gene*** | ***test*** | ***Clinically identified variant*** |  | ***VAF*** | | | |
| --- | --- | --- | --- | --- | --- | --- | --- | --- |
|  |  |  |  | ***variant type*** | ***cfDNA 1*** | ***cfDNA 2*** | ***sphDNA*** | ***FFPE*** |
| ASC6 | *TP53* | Somatic | p.C242Afs*5 | Frameshift | 0.59 | NT | NT | NT |
|  | *NF1* | Somatic | p.Y49* | Nonsense | 0.61 | NT | NT | NT |
| ASC9 | *TP53* | Somatic | p.R273C | Missense LOF | 0.71 | 0.73 | 0.09 | 0.79 |
| ASC11 | *BRCA1* | Somatic | p.K1606Lfs*18 | Frameshift | 0.34 | 0.31 | 0.06 | NT |
| ASC12 | *TP53* | Somatic | p.S241Ffs*23 | Frameshift | 0.77 | 0.53 | NT | NT |
|  | *BRCA2*  *RB1* | Somatic  Somatic | p.Y748*  p.L468* | Nonsense  Nonsense | 0.79  0.91 | 0.58  0.52 | NT  NT | NT  NT |
| ASC17 | *BRCA2* | Somatic | p.W2169* | Nonsense | 0.31 | NT | NT | 0.23 |
|  | *RAD51D* | Genomic | p.C9S (VUS) | Missense (Amb) | 0.57 | NT | NT | 0.59 |
| ASC20 | *BRCA1* | Genomic | p.L785* | Nonsense | 0.82 | 0.74 | 0.84 | NT |
|  | *TP53* | Somatic | p.P278A | Missense LOF | 0.75 | 0.55 | 0.80 | NT |
| ASC23 | *TP53* | Somatic | p.R196* | Nonsense | 0 | 0 | 0 | 0.76 |
|  | *PIK3CA* | Somatic | p.E545K | Missense GOF | 0.2324 | 0.4364 | 0.0792 | 0.27 |
| ASC28 | *PALB2* | Unknown | p.S1102R (VUS) | Missense LOF | 0.48 | 0.50 | 0.50 | 0.52 |
| ASC30 | *TP53* | Somatic | c.672+1G>T | Splice site | 0.88 | NT | NT | 0.74 |
| ASC36 | *BRCA1* | Genomic | p.V1736A | Missense LOF | 0.65 | 0.68 | NT | NT |
|  | *BRCA2* | Genomic | p.D3073G | Missense LOF | 0.44 | 0.44 | NT | NT |
| ASC38 | *BRCA1* | Genomic | p.L392GInfs*5 | Frameshift | 0.75 | NT | NT | NT |
|  | *TP53* | Somatic | p. Y236C | Missense | 0.61 | NT | NT | NT |

NT, not tested; LOF, loss of function; GOF, gain of function; Amb, ambiguous effect

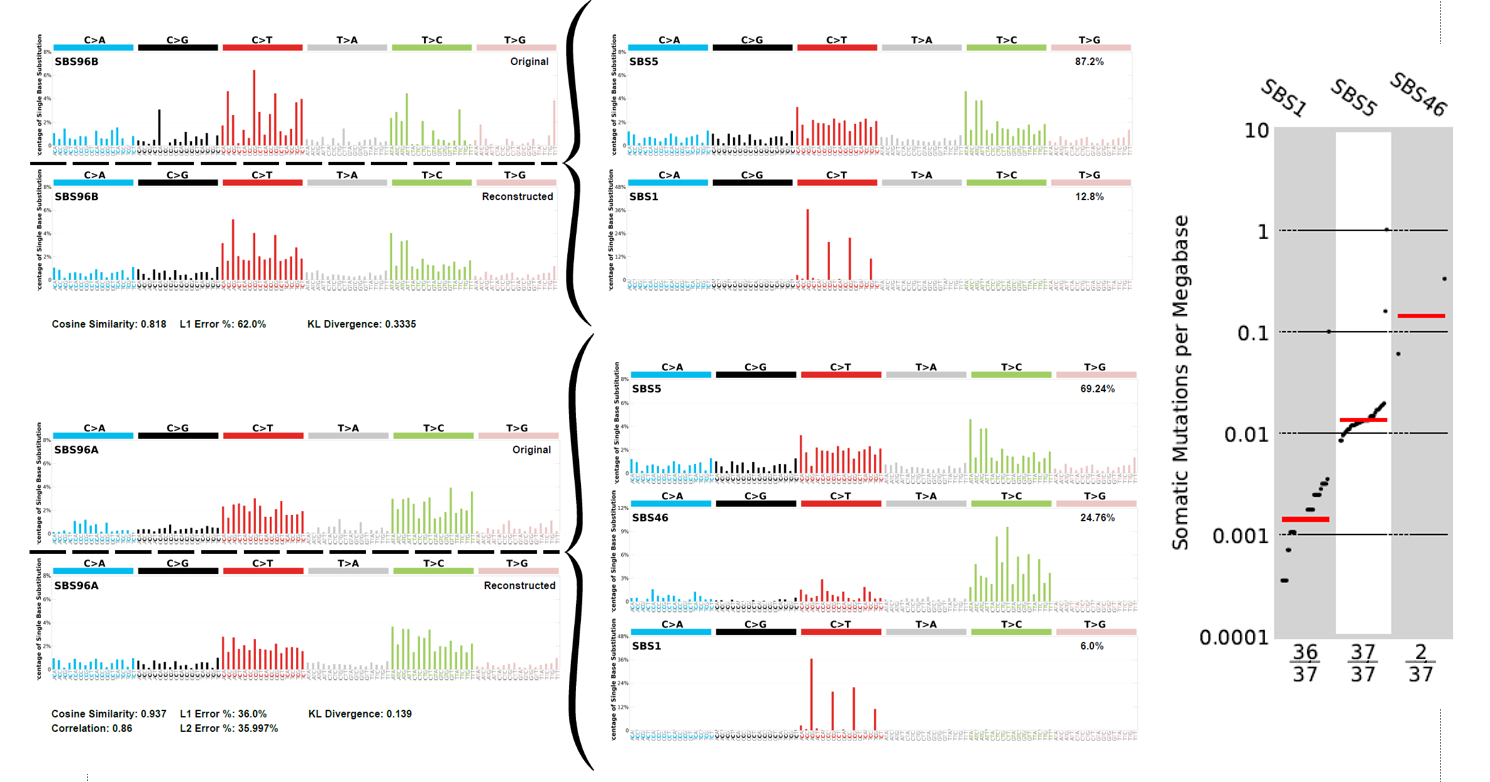

**Supplementary Figure 4.** *COSMIC Single Base Substitution Signatures assigned to samples*

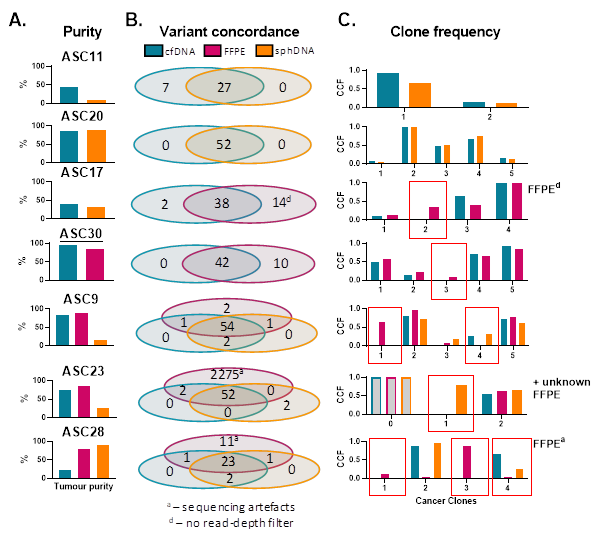

***Supplementary Figure 5.*** *Variant identification and clonal representation across sample types. Concordance in somatic variants reported in cfDNA (blue), sphDNA (orange) and FFPE (maroon) (A). Cancer cell fraction (CCF) in clonal clusters identified by PyClone-VI (B). Red boxes outline clones not common across sample set. CCF, cancer cell fraction; cfDNA, cell-free DNA; sphDNA, DNA from ascites-derived cell spheroids; FFPE, DNA from formaldehyde-fixed paraffin embedded tumour biopsy samples. For ASC30 (underlined) FFPE and cfDNA were collected at matched timepoint, otherwise, all FFPE samples were collected independently. All cfDNA and sphDNA collected at matched timepoint.*

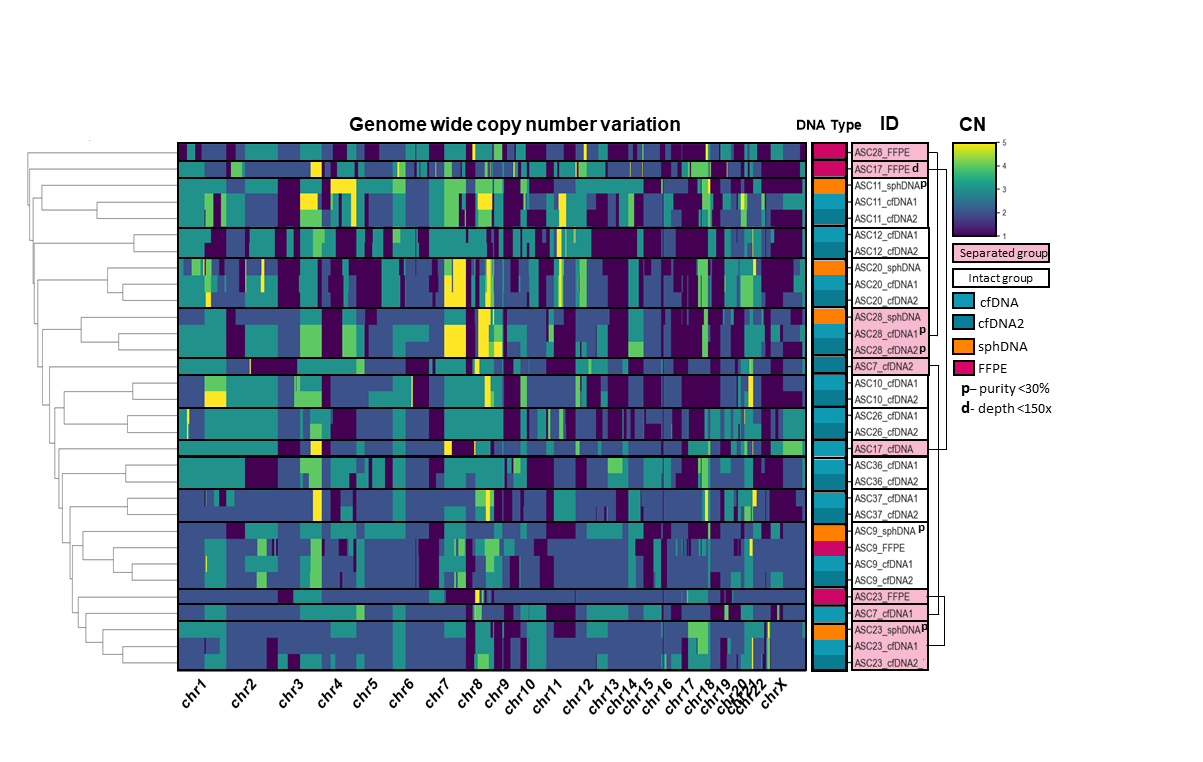

**Supplementary Figure 6.** *Copy number consensus between samples.* Copy number profiles generated by CNVKit.

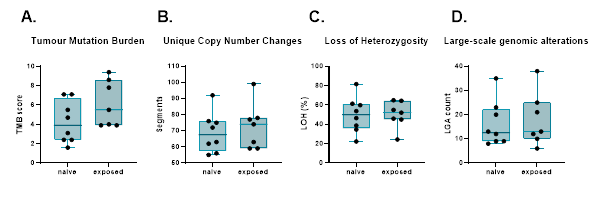

***Supplementary Figure 7.*** *Tumour mutation burden (TMB) (A), copy number segmentation (B) and loss of heterozygosity (C) in initial cell-free DNA samples, separated by preceding chemotherapy exposure, analysed by Mann-Whitney test (no significance).*

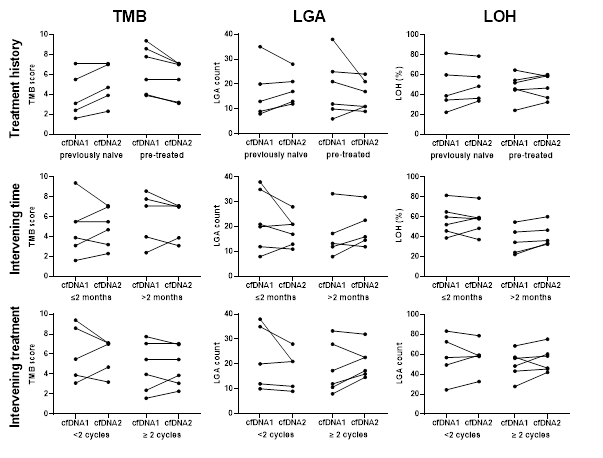

**Supplementary Figure 8.** *Tumour mutation burden large-scale genomic alterations and loss of heterozygosity in sequential ascites samples, separated into individuals with disparate treatment history, intervening time length and amount of intervening treatment.*
